# A case-cluster of aseptic meningitis associated with a newly identified recombinant echovirus6/CoxsackievirusB1 enterovirus

**DOI:** 10.1101/2024.12.11.24318716

**Authors:** Kartikeya Cherabuddi, Massimiliano S. Tagliamonte, John A. Lednicky, David A. Ostrov, Tracey L. Moquin, Vishal Kaushik Thoomkuntla, Brian Bourgeois, Nicole M. Iovine, Paul D. Myers, Marco Salemi, J. Glenn Morris

## Abstract

A daycare teacher presented to a local Emergency Department (ED) with complaints of headache, neck stiffness, and fever. Preliminary analysis of cerebral spinal fluid (CSF) raised concerns about meningococcal meningitis and prompted notification of the county health department, which then notified the daycare. Ten children presented to a University referral hospital for evaluation, four of whom were febrile. CSF from the teacher and nasal swabs from the febrile children were all RT-PCR positive for enterovirus. A novel recombinant enterovirus was cultured from the teacher’s CSF and from two of the nasal swabs. The amino-terminal portion of the recombinant virus was derived from an Echovirus E6 with the carboxy-terminal portion originating from a Coxsackievirus B1; recombinant segments were most closely related to similar segments from strains isolated in France. Recombination occurred within the C-2 gene which encodes a multifunctional protein that functions as an RNA-stimulated ATPase associated with virus replication and virion morphogenesis. Structural modeling predicted that the recombinant protein was capable of forming hexameric and heptameric assemblies. Our data highlight the ongoing potential for recombination among enteroviruses groups, leading to modifications within viral proteins which may impact virulence.

**Brief Article Summary:** After a daycare teacher was diagnosed with meningitis, ten children were referred by the county health department to a University Hospital for evaluation. Cerebral Spinal Fluid (CSF) from the teacher and nasal swabs from symptomatic children were positive for a novel recombinant enterovirus, with the amino-terminal portion of the virus derived from Echovirus E6 and the carboxy-terminal portion originating from a Coxsackievirus B1.

## INTRODUCTION

The genus *Enterovirus* consists of small single-stranded positive-sense RNA viruses, which are generally transmitted via a respiratory route or through the gastrointestinal tract. Enteroviruses are within the *Picornavirus* family and are responsible for a wide range of illnesses in humans, including upper respiratory infections, meningitis/encephalitis, myopericarditis, and poliomyelitis. Enteroviruses were previously classified into four major groups: polioviruses, Coxsackie A viruses, Coxsackie B viruses, and echoviruses. Current nomenclature is based on genetic characteristics, with identification of 15 enterovirus/rhinovirus species, which may contain both echovirus and Coxsackievirus serotypes [1–3]. RNA recombination occurs frequently within enteroviral groups and is a major driving force in the evolution of the species [4–6]. We report here a cluster of enterovirus cases associated with a childcare facility; the virus responsible for the outbreak was a novel recombinant of echovirus 6 and coxsackievirus B1.

## METHODS

Clinical and epidemiologic data were obtained through the University referral hospital Infection Control Program and the County Department of Health. Cerebral spinal fluid (CSF) and nasal swab samples were tested by PCR/RT-PCR via the BioFire meningitis/encephalitis panel and the respiratory 2.1 panel (bioMérieux, Durham, North Carolina, USA), respectively. With this platform enteroviruses are reported as “rhinovirus/enterovirus.” The study was reviewed by the University of Florida IRB, which waived ethical approval for the work; the index case provided written consent for inclusion of data in the report.

### Viral culture/sequencing

The following cell lines obtained from the American Type Culture Collection (ATCC) were chosen for virus isolation attempts, as previously described [7]: A549 (CCL-185), LLC-MK2 (CCL-7), and MRC-5 (CCL-171). All the cell lines were propagated as monolayers at 37°C and 5% CO_2_ in advanced Dulbecco’s modified Eagle’s medium (aDMEM, Invitrogen, Carlsbad, CA) supplemented with 10% heat-inactivated, gamma-irradiated, low-antibody fetal bovine serum (FBS) (GE Healthcare Life Sciences, Pittsburgh, PA, Cat#: SH30070.03IH), 2 mM L-Alanyl-L-Glutamine (GlutaMAX, Invitrogen Corp.) and antibiotics (PSN; 50 μg/ml penicillin, 50 μg/ml streptomycin, 100 μg/ml neomycin [Invitrogen Corp.]). RNA was extracted from virions in the spent cell media of A549 cells using a QIAamp Viral RNA Mini Kit (Qiagen, Valencia, CA, USA) according to the manufacturer’s protocol. A cDNA library was generated using a NEBNext Ultra RNA Library Prep kit (New England Biolabs, Ipswich, Massachusetts, USA), and the library sequenced on an Illumina NextSeq 1000 sequencer. De novo assembly of the paired-end reads was performed using MEGAHIT v1.1.4 [8].

### Phylogenetic studies

Using BLAST, matches to the full genome sequences were obtained and filtered based on ≥70% percent identity. Non-coding UTRs were trimmed out as they were missing from many of the sequences, keeping only the polyprotein segment for downstream analyses. The alignment was subsampled by country taking into consideration genetic diversity and temporal sampling using TARDIS [9,10] keeping a maximum of five sequences per country. To the dataset we added sequences that showed close genetic relation to the major fragments at a preliminary analysis (OR840837.1, OR840843.1, OR840844.1), and four more distantly related strains as outgroup (JX982254.1, MF678294.1, JN542510.1, JX174176.1). The final dataset resulted in 133 isolates, including isolates from the present study. Recombination analyses and phylogenetic signal assessment were performed as previously described [11,12].

Phylogenetic analysis was performed by inferring a neighbor-joining tree using Mega using the best fitting nucleotide substitution model and 1000 bootstrap replicates [13]. A relaxed molecular clock for each recombinant segment was calibrated using dated tips and the Bayesian coalescent framework implemented in BEAST [14] by running a Markov Chain Monte Carlo for 50 million generations with a sampling frequency of 50,000. Final maximum clade credibility tree was calculated with TreAnnotator and displayed in FigTree (http://tree.bio.ed.ac.uk/software/figtree/). E.S.S of all estimated parameters were > 200, showing proper mixing of the Markov chain.

### Modeling of Protein Structure

Translated virus sequences were used as input for structural prediction using AlphaFold 3 [15]. The resulting predicted structures were aligned and superimposed using Secondary Structure Matching (SSM) in COOT [16]. Structural analyses, including atomic distance measurements, were conducted, and visual representations were generated with PyMol [17].

## RESULTS

### Outbreak

A teacher (aged 25-30 years) at a community daycare presented to a local Emergency Department (ED) with complaints of severe headache, neck stiffness, and high-grade fevers; the teacher also reported photophobia, nausea, and an episode of vomiting. A lumbar puncture was performed, and cerebral spinal fluid (CSF) showed a WBC count of >500 cells/ml with 98% PMNs. A Gram stain was read initially as showing Gram-negative cocci in pairs; on a repeat evaluation it was determined that this was an artifact and the Gram stain was officially reported as being negative. However, because of the initial concerns that this represented meningococcal or other bacterial meningitis, the patient was started empirically on vancomycin and ceftriaxone.

In the setting of a possible meningococcal meningitis case the ED notified the county health department, and per protocol the health department contacted the daycare and advised that if parents had concerns about possible exposure they should bring their children to a University referral hospital for further evaluation. Ten children came to the hospital for evaluation, both daycare attendees and their siblings; because the daycare had been closed for the holidays, exposures ranged from 5-8 days prior to the hospital visit. Four children were symptomatic, three of whom underwent lumbar puncture; symptoms and laboratory results are shown in Table 1. Based on the report of the teacher’s CSF pleocytosis, all four were started empirically on vancomycin and ceftriaxone. Potential adverse reactions to the antimicrobial therapy were noted in three children, resulting in administration of administration of diphenhydramine and, in one case, methylprednisolone and epinephrine.

**Table 1:**
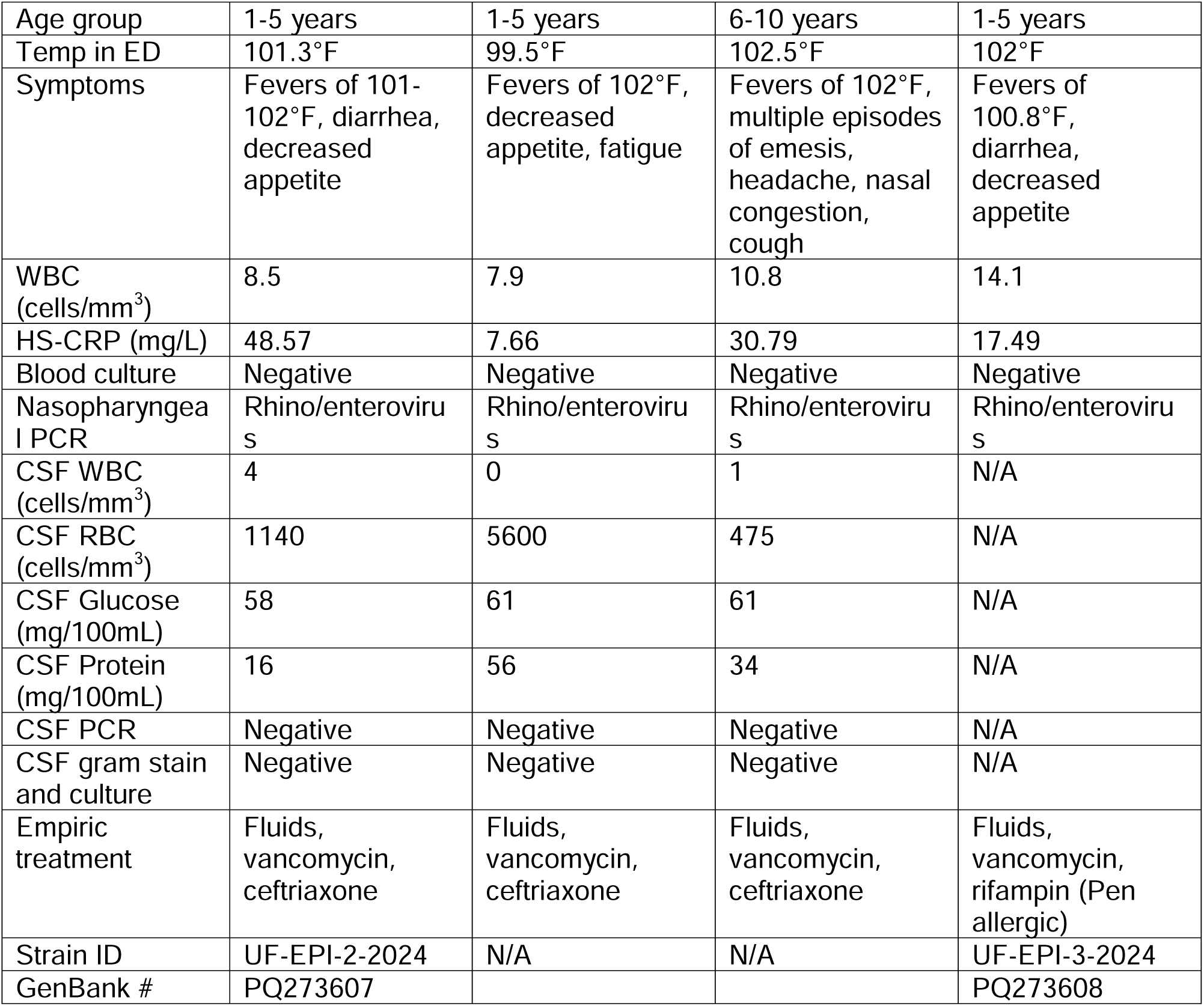
Clinical data, children with positive nasopharyngeal swabs for enterovirus.

|  |  |  |  |  |
| --- | --- | --- | --- | --- |
| Age group | 1-5 years | 1-5 years | 6-10 years | 1-5 years |
| Temp in ED | 101.3°F | 99.5°F | 102.5°F | 102°F |
| Symptoms | Fevers of 101-102°F, diarrhea, decreased appetite | Fevers of 102°F, decreased appetite, fatigue | Fevers of 102°F, multiple episodes of emesis, headache, nasal congestion, cough | Fevers of 100.8°F, diarrhea, decreased appetite |
| WBC (cells/mm<sup>3</sup>) | 8.5 | 7.9 | 10.8 | 14.1 |
| HS-CRP (mg/L) | 48.57 | 7.66 | 30.79 | 17.49 |
| Blood culture | Negative | Negative | Negative | Negative |
| Nasopharyngeal PCR | Rhino/enteroviruses | Rhino/enteroviruses | Rhino/enteroviruses | Rhino/enteroviruses |
| CSF WBC (cells/mm<sup>3</sup>) | 4 | 0 | 1 | N/A |
| CSF RBC (cells/mm<sup>3</sup>) | 1140 | 5600 | 475 | N/A |
| CSF Glucose (mg/100mL) | 58 | 61 | 61 | N/A |
| CSF Protein (mg/100mL) | 16 | 56 | 34 | N/A |
| CSF PCR | Negative | Negative | Negative | N/A |
| CSF gram stain and culture | Negative | Negative | Negative | N/A |
| Empiric treatment | Fluids, vancomycin, ceftriaxone | Fluids, vancomycin, ceftriaxone | Fluids, vancomycin, ceftriaxone | Fluids, vancomycin, rifampin (Pen allergic) |
| Strain ID | UF-EPI-2-2024 | N/A | N/A | UF-EPI-3-2024 |
| GenBank # | PQ273607 |  |  | PQ273608 |

CSF from the teacher was subsequently reported to be culture-negative for bacterial pathogens, with, as noted, a negative Gram stain on repeat evaluation. CSF from the teacher and nasal swabs from all four febrile children were tested at the University referral hospital and found to be RT-PCR-positive for “rhinovirus/enterovirus.” Based on these results, antimicrobial therapy was discontinued in all cases. All recovered without further sequelae.

### Virus culture

CSF and nasal swab samples were sent to the University of Florida Emerging Pathogens Institute for virus culture. Virions in the CSF sample from the teacher and nasal swabs from two of the children caused virus-induced cytopathic effects (CPE) by three days post inoculation (dpi) of A549 cells, and 4 dpi of LLC-MK2 cells. The CPE consisted of swelling of the cells, followed by their detachment from the growing surface. By 5 dpi, much of A549 cell culture had detached from the growing surface, whereas only about 30% of the LLC-MK2 cells displayed CPE. No CPE were detected in MRC-5 cells. This pattern was somewhat unexpected: our previous experience with other echovirus strains has been that the relative magnitude of the CPE responses were LLC-MK2 = MRC5 > A549 cells. No CPE were observed in mock-inoculated cells maintained in parallel. All three virus strains cultured were sequenced and sequences submitted to GenBank (GenBank numbers: adult case, PP873675; children [Table 1] PQ273607, PQ273608)

### Phylogenetic analysis

The three clinical virus strains isolated from culture had essentially identical sequences, consistent with clonal spread of the virus within the daycare/home setting. On further analysis, we found that the amino-terminal portion of the virus showed homology with Echovirus E6, while the carboxy-terminal portion appears to have originated from a Coxsackievirus B1, with a junction in the non-structural gene 2C (Figure 1). Figure 2A shows the neighbor joining tree inferred for the VP4-VP2-VP3-VP1-2A-2B concatemer of our patients’ virus clustering with Echovirus E6, while Figure 2B shows clustering of the 3A-3B-3C-3D concatemer with Coxsackievirus 1B. Figures 2C and 2D show maximum clade credibility (MCC) trees for these two segments. For both, segments cluster most closely to segments from French strains, suggesting that the recombination event occurred in France, possibly in or around 2019. There is not clear evidence of clustering of CSF isolates (shown in red in both sets of trees) for either concatemer.

**Figure 1.**
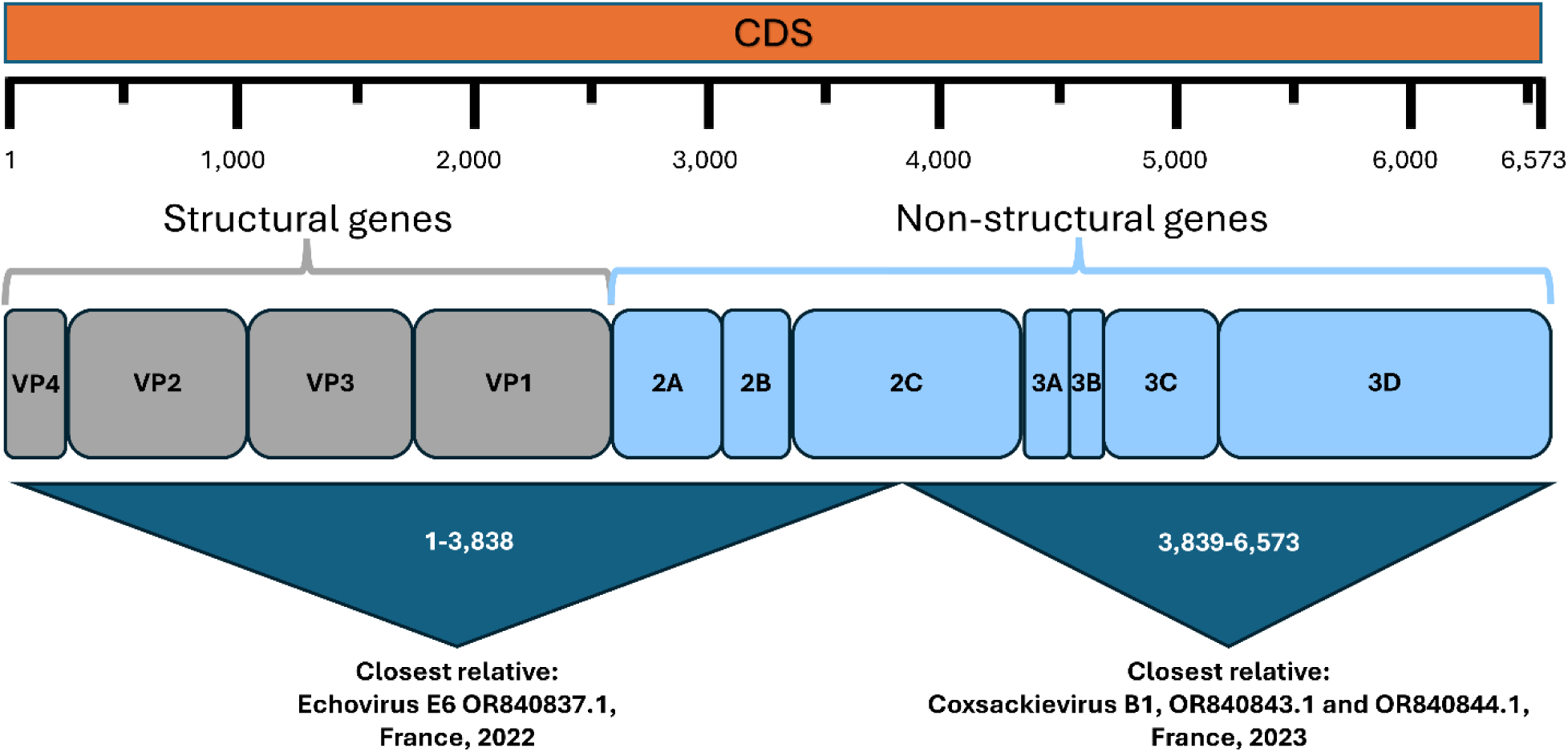
Schematics of the CDS genomic segment, with gene structure and summary of their ancestry from the phylogenetic analysis. Coordinates are based on the UF-EPI-1-2024 strain from the initial adult patient.

**Figure 2.**
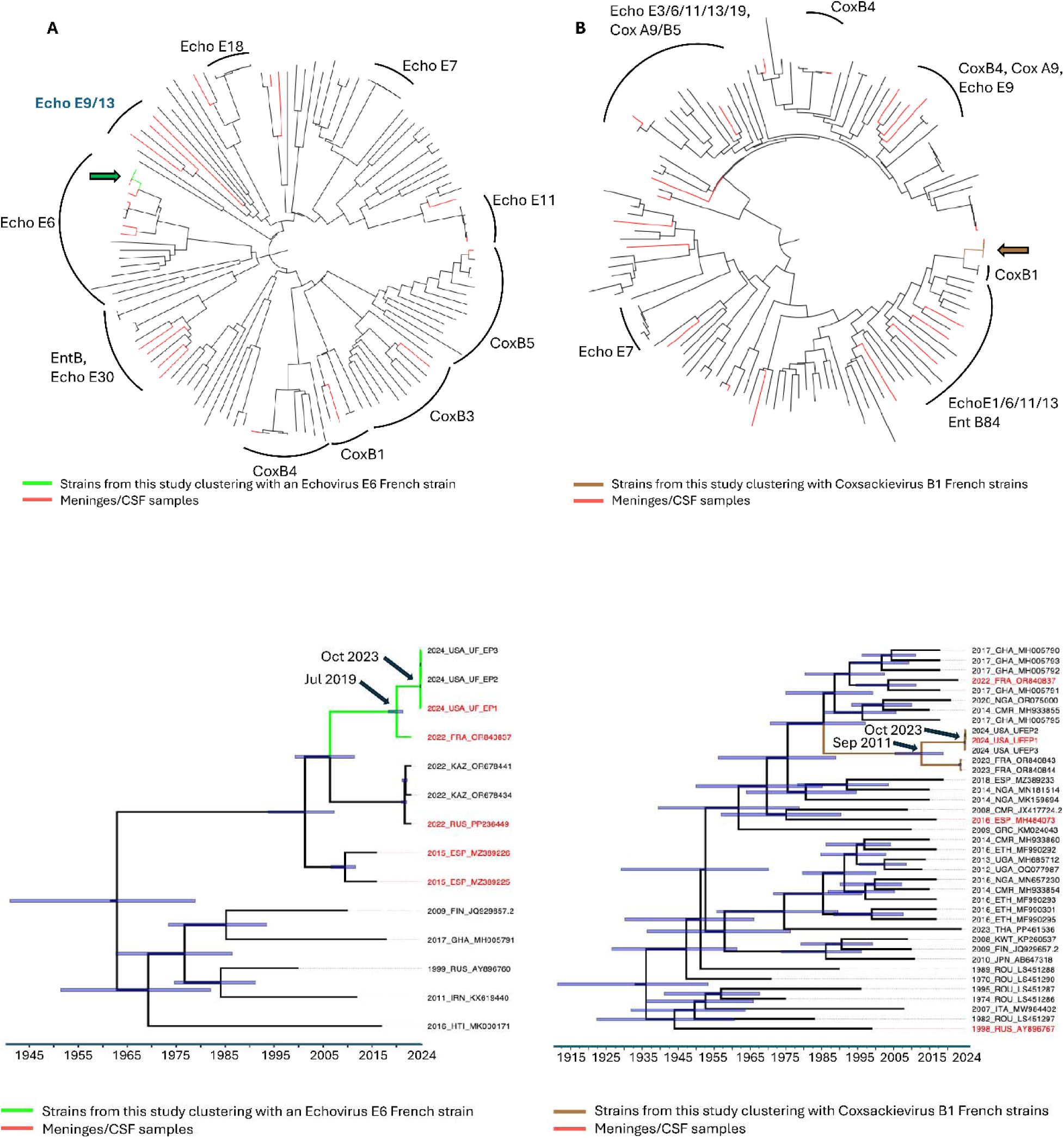
Panels A/B: Neighbor-joining (NJ) trees inferred from the two major non recombinant segments of the full genome sequences. Branch lengths are drawn proportional to nucleotide substitutions per site according to the scale at the bottom. Red terminal branches indicate sequences sampled from meninges and/or CSF. Arcs indicate strains belonging to well supported clades (bootstrap >90%). Echo=Echovirus, Ent=Enterovirus, Cox= Coxsackievirus. **Panel A.** NJ tree inferred from the VP2-VP3-VP1-2A-2B-2C non recombinant concatemer. Strains isolated in this study (green branches) are indicated by a green arrow. **Panel B.** NJ tree inferred from the 3A-3C-3D non recombinant concatemer. Strains isolated in this study (brown branches) are indicated by a brown arrow. **Panels C/D: Bayesian clock-like phylogenies inferred from the two major non recombinant segment of the full genome sequences.** For each tree includes a subset of sequences most closely related to the strains isolated in this study. Branch lengths are scaled in time according to the scale at the bottom. Red labels indicate sequences sampled from meninges and/or CSF. Horizontal bars represent the 95%HPD interval of estimated dates for highly supported (P>0.95) internal nodes. **Panel C.** Maximum clade credibility (MCC) tree inferred from the VP2-VP3-VP1-2A-2B-2C non recombinant concatemer of Echovirus 6 strains. Green branches: monophyletic clade clustering the new USA isolates with an Echovirus 6 French strain. **Panel D.** Maximum clade credibility (MCC) tree inferred from the 3A-3C-3D non recombinant concatemer of Echovirus and Coxsackie virus strains. Brown branches: monophyletic clade clustering the new USA isolates with two Coxsackievirus B1 French strains highlighted in green.

### Modeling of Protein Structure

AlphaFold 3 was employed to generate three-dimensional structural models of the 2C protein from the French Echovirus 6 and Coxsackievirus B1 strains, and the recombinant Florida viruses. The Florida viruses were found to contain a chimeric 2C protein, with the amino-terminal portion derived from Echovirus 6 and the carboxy-terminal region originating from Coxsackievirus B1. Mutational analysis revealed that changes in the Florida viruses were distributed across both the amino- and carboxy-terminal regions of the 2C protein (Figure 3). Structural predictions by AlphaFold 3 indicated a rearrangement of the first α-helix in the 2C protein of the Florida viruses, introducing a break near the mutated position 23. This structural rearrangement, together with additional alterations in the 2C protein, was predicted to influence subunit assembly.

**Figure 3.**
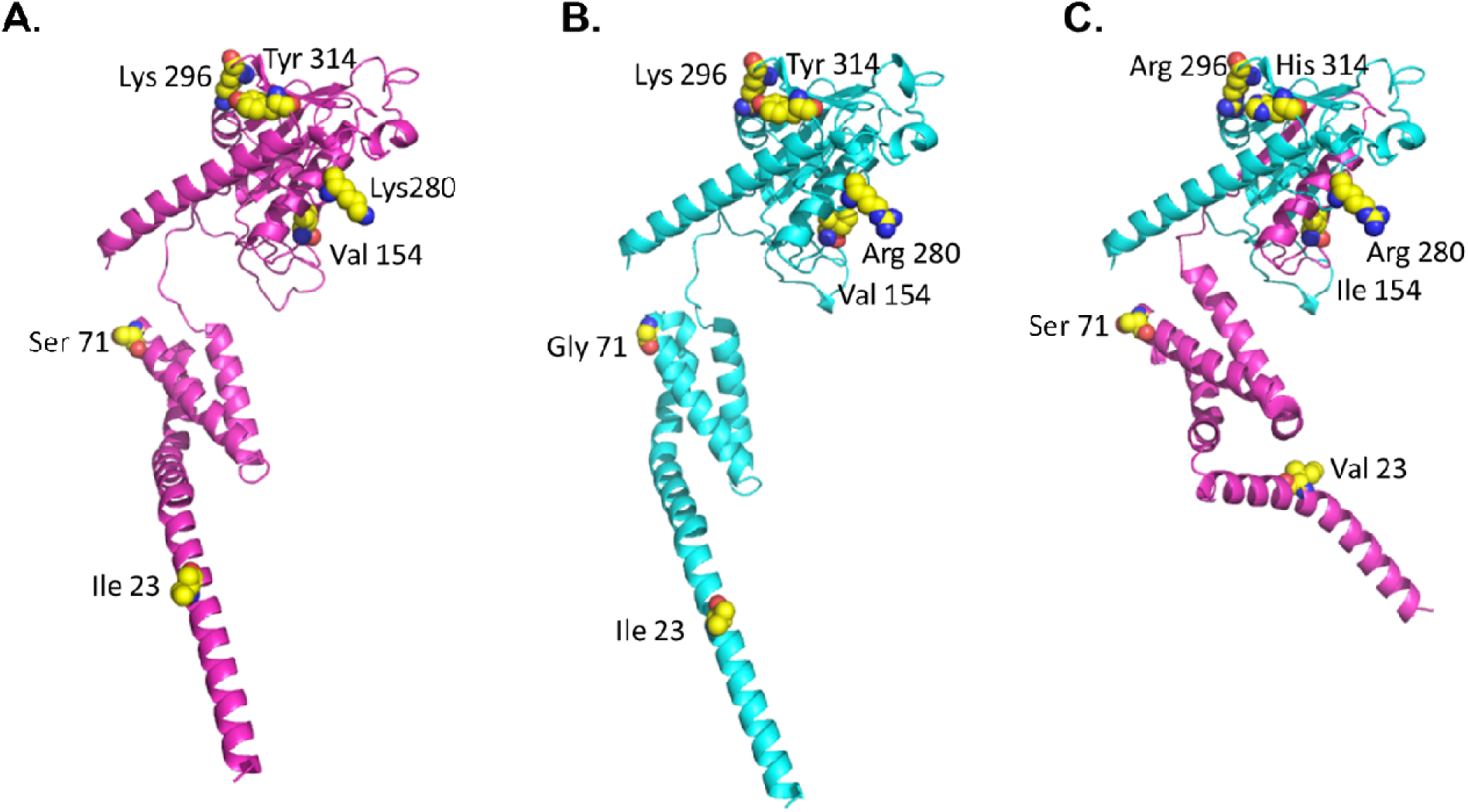
Recombination and mutation result in the emergence of novel viruses identified in Florida. **Panel A** shows a ribbon diagram depicting the structural prediction of an Echovirus E6 polypeptide (2022_FRA_OR840837.1) shown in magenta. **Panel B** illustrates a ribbon diagram of the structural prediction for a Coxsackie virus polypeptide (2023_FRA_OR840844.1) shown in cyan. **Panel C** displays a ribbon diagram of the predicted structure for a recombinant polypeptide, integrating sequences from Echovirus E6 (magenta) and Coxsackie virus (cyan)(2024_USA_UF-EP1-3-2024_FL). The residues encoded by the 5’ and 3’ recombinant fragments come from the separate parental strains (magenta for Echovirus E6, cyan for Coxackie virus). Positions mutated following recombination are shown as spheres, yellow for carbon, blue for nitrogen, red for oxygen.

AlphaFold 3 further predicted distinct oligomerization patterns for the 2C protein. Specifically, Echovirus 6 2C was modeled as forming a hexameric assembly (Figure 4A and E), consistent with previous experimental observations [18]. In contrast, Coxsackievirus B1 2C was predicted to form heptameric assemblies (Figure 4B and F). Interestingly, the recombinant Florida viruses were predicted to adopt both hexameric (Figure 4C and G) and heptameric assemblies (Figure 2D and 2H), suggesting that recombination and mutation enable these viruses to mimic molecular assemblies characteristic of both parental viruses. Arg at position 280 of the Florida viruses has the potential to mediate intermolecular contacts with neighboring chains in the hexameric and heptameric assemblies (Figure 5). These findings highlight the structural plasticity imparted by recombination and mutation in the evolution of viral proteins.

**Figure 4.**
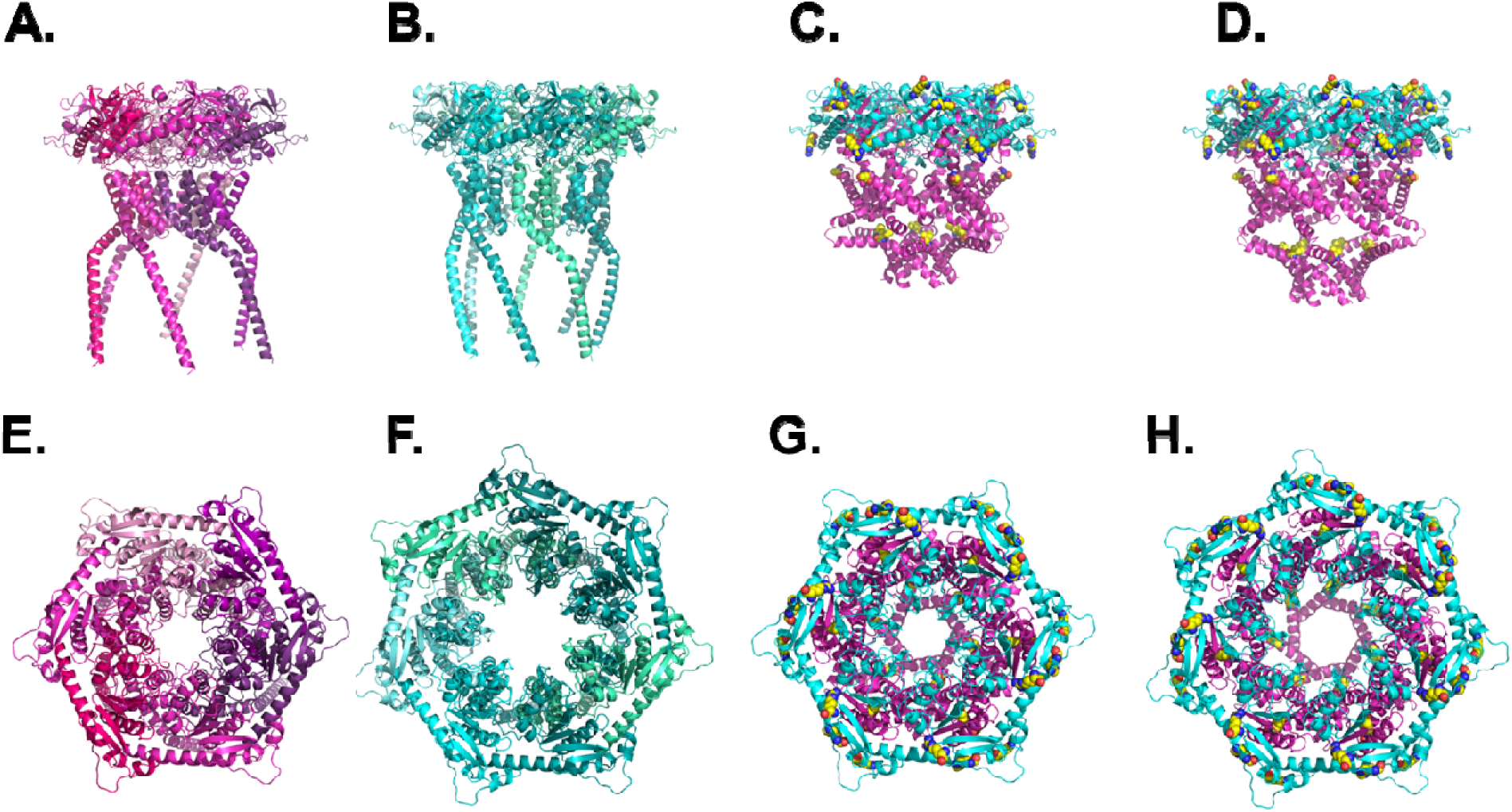
Recombination and mutation in Coxsackie virus are predicted to impact subunit assembly. AlphaFold 3 predictions indicated that the Echovirus 2C polypeptide forms hexamers, whereas the Coxsackie virus 2C polypeptide forms heptamers. The recombinant mutated viruses identified in Florida express 2C polypeptides predicted to form hexamers and heptamers. **Panels A** and **B** display side views of the Echovirus hexamer and Coxsackie virus heptamer, respectively. **Panels C** and **D** show side views of the recombinant mutated virus predicted to form a hexameric and heptameric arrangement, respectively, highlighting that the amino-terminal portion of the recombinant virus is derived from Echovirus E6 (shown in magenta), while the carboxy-terminal portion originates from Coxsackie virus (cyan). **Panels E** and **F** display top views of the Echovirus hexamer and Coxsackie virus heptamer, respectively. **Panels G** shows the top view of the recombinant mutated virus predicted to form a hexameric arrangement. **Panels H** shows the top view of the recombinant mutated virus predicted to form a heptameric arrangement.

**Figure 5.**
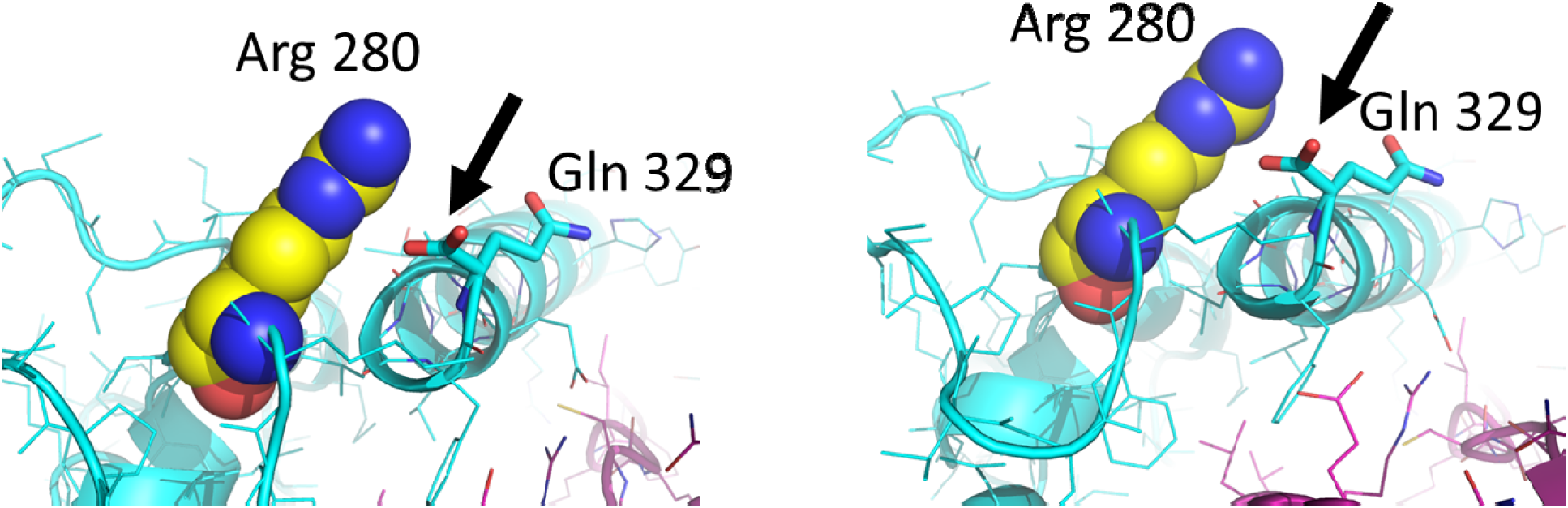
Mutated positions in the recombinant virus impact intermolecular interactions between hexameric and heptameric subunits. **Panel A** shows that Arg at position 280 forms an intermolecular contact with neighboring chains by interacting with the terminal carboxyl group (arrow) of Gln at position 329 in the hexameric assembly. **Panel B** shows that Arg 280 mediates intermolecular contact with neighboring chains in the heptameric assembly by contacting the carboxyl group at position 329 (arrow).

## DISCUSSION

Clinically, this outbreak is consistent with what has been previously described in association with enteroviruses. Our data document transmission of the virus within a daycare setting and within households, in keeping with the “transmission clusters” reported from Europe and other parts of the world [2,19]. While the CSF neutrophil pleocytosis seen in our adult patient raised concerns about bacterial infection, in this instance the positive RT-PCR and virus culture for enterovirus and negative bacterial culture and repeat Gram-stain are consistent with an enteroviral etiology, and underscore both the possibility of an initial PMN response in a viral infection and the importance of PCR/RT-PCR and culture in identification of an etiologic agent. This report also raises the issue of shortage of well-trained clinical laboratory workers, including microbiology technologists [20], limiting performance and interpretation of basic laboratory tests such as Gram strains, especially after day-shift hours. Given the reported adverse reactions to antimicrobial therapy seen in three of the children in the outbreak, necessitating administration of solumedrol and epinephrine in one case; stress in parents with concern for meningitis exposure to their children; and the strain on a pediatric ED receiving multiple concerned families in a short period of time, the availability of rapid RT-PCR testing of CSF and nasopharyngeal swabs for agents such as enterovirus is of particular importance in an ED setting.

Both echovirus 6 and Coxsackievirus B1 have been placed within the enterovirus species *Enterovirus betacoxackie* [3], previously referred to as EV-B. Echovirus 6 is well recognized as one of the most common causes of aseptic meningitis, and has been implicated in meningitis outbreaks [2,19]. Coxsackievirus B1 has also been linked with aseptic meningitis, as well as myocarditis, pancreatitis, hand, foot, and mouth disease, and has been implicated as a possible trigger for the development of type 1 diabetes [21]. The recombination site for the recombinant virus which we identified in this outbreak was within the nonstructural protein 2C, which encodes a RNA-stimulated ATPase [22] associated with virus uncoating, host cell membrane rearrangements, RNA replication, encapsidation, morphogenesis, ATPase, helicase, and chaperoning activities.

The structural changes in the 2C protein of the Florida viruses, particularly the rearrangement of the first α-helix and the ability to form both hexameric and heptameric assemblies, may have significant functional and evolutionary implications. The α-helix break near position 23, along with mutations in the amino- and carboxy-terminal regions, likely alters the stability and dynamics of the 2C protein, potentially influencing its role in viral replication and assembly. The capacity to adopt multiple oligomeric states could enhance the flexibility of the Florida viruses in adapting to different host environments or cellular conditions, providing a potential selective advantage. Moreover, the ability to emulate structural features of both Echovirus 6 and Coxsackievirus B1 suggests that recombination and mutation have conferred novel functional properties, which may impact virus-host interactions, immune evasion, or pathogenesis. These structural modifications could thus play a critical role in the emergence and dissemination of our Florida viruses, making them a unique model for studying the effects of structural diversity on viral fitness and evolution.

In summary, this report highlights the potential for enterovirus case clusters and the need for rapid diagnostics capable of identifying enterovirus infections. While the propensity for enteroviruses to recombine is well recognized, the 2C gene is reported to have a low frequency of recombination, and recombination between groups (in this case, echovirus and Coxsackievirus) is not well described. While we do not have a complete understanding of the significance of this particular recombination and its potential for increasing virulence, the adult case in this outbreak was, indeed, severe, with the Echovirus 6 gene segment most closely related to the segment in our Florida strains also coming from a strain isolated from CSF (Figure 2A and 2C). Given that viruses associated with aseptic meningitis are seldom cultured or sequenced, we have no way of estimating the degree of spread of this virus. The recognition that such recombinants are continuing to emerge [5,6] underscores the need for ongoing surveillance of enteroviral infections (with sequence analysis), particularly in instances with more severe illness or case clusters.

## Data Availability

All data produced in the present work are contained in the manuscript or have been submitted to GenBank

## Acknowledgments

Bioinformatic analyses were in part performed by the UF-ICBR Bioinformatics Core (RRID:SCR_019120).

## Presentations

Data have not been previously presented in abstract form at meetings.

## Corresponding author

J. Glenn Morris, Jr., MD, MPH&TM

Professor and Director, Emerging Pathogens Institute

University of Florida

2055 Mowry Road, Gainesville, FL 32610-0009

Alternative corresponding author: Dr. John Lednicky

## REFERENCES

1. Wright WF, Pinto CN, Palisoc K, Baghli S. Viral (aseptic) meningitis: A review. J. Neuro. Sci. 2019;398:176–183

2. Monge S, Benschop K, Soetens L, Pijnacker R, Hahne S, Wallinga J, Duizer E. Echovirus type 6 transmission clusters and the role of environmental surveillance in early warning, the Netherlands, 2007-2016. Euro Surveill. 2018;23:pli=1800288.

3. ICTV. Virus taxonomy: the ICTV report on virus classification and taxon nomenclature. https://ictv.global/report/chapter/picornaviridae/picornaviridae/enterovirus. Accessed 11/21/2024.

4. Muslin C, Kain AN, Bessaud M, Blondel B, Delpeyroux F. Recombination in enteroviruses, a multi-step modular evolutionary process. Viruses 2019;11:859; doi:10.3390/vii090859

5. Gong Y-N, Yang S-L, Chen Y-C, et al. Novel intertypic recombination of Echovirus 11 in the Enterovirus species B. J Med Virol 2024;96:e29323. 10.1002/jmv.29323.

6. Wang X, Cun J, Li S, et al. Genotype F of Echovirus 25 with multiple recombination pattern have been persistently and extensively circulating in Chinese mainland. Scientific Reports 2024;24:3212 10.1038/s41598-024-535132.

7. Pan M, Bonny TS, Loeb J, Jiang X, Lednicky JA, Eiguren-Fernandez A, Hering S, Fan ZH, Wu C. 2017. Collection of Viable Aerosolized Influenza Virus and Other Respiratory Viruses in a Student Health Care Center through Water-Based Condensation Growth. mSphere 2:10.1128/msphere.00251-17. https://doi.org/10.1128/msphere.00251-17

8. DeRuyter, E.; Subramaniam, K.; Wisely, S.M.; Morris, J.G., Jr.; Lednicky, J.A. A Novel Jeilongvirus from Florida, USA, Has a Broad Host Cell Tropism Including Human and Non-Human Primate Cells. Pathogens 2024, 13, 831. 10.3390/pathogens13100831

9. Altschul SF, Gish W, Miller W, Myers EW, Lipman DJ. Basic local alignment search tool. J Mol Biol. 1990;215(3):403–10.

10. Marini S, Mavian C, Riva A, Prosperi M, Salemi M, Rife Magalis B. Optimizing viral genome subsampling by genetic diversity and temporal distribution (TARDiS) for phylogenetics. Bioinformatics. 2022;38(3):856–60.

11. Lednicky JA, Tagliamonte MS, White SK, Elbadry MA, Alam MM, Stephenson CJ, Bonny TS, Loeb JC, Telisma T, Chavannes S, Ostrov DA, Mavian C, Beau De Rochars VM, Salemi M, Morris JG Jr. Independent infections of porcine deltacoronavirus among Haitian children. Nature. 2021 Dec;600(7887):133-137. doi: 10.1038/s41586-021-04111-z. Epub 2021 Nov 17. PMID: 34789872; PMCID: PMC8636265.

12. Lednicky JA, Tagliamonte MS, White SK, Blohm GM, Alam MM, Iovine NM, Salemi M, Mavian C, Morris JG. Isolation of a Novel Recombinant Canine Coronavirus From a Visitor to Haiti: Further Evidence of Transmission of Coronaviruses of Zoonotic Origin to Humans. Clin Infect Dis. 2022 Aug 24;75(1):e1184–e1187. doi: 10.1093/cid/ciab924. PMID: 34718467; PMCID: PMC9402678.

13. Kumar S, Stecher G, Li M, Knyaz C, Tamura K. MEGA X: Molecular Evolutionary Genetics Analysis Across Computing Platforms. Molecular biology and evolution. 2018;35(6).

14. Suchard MA, Lemey P, Baele G, Ayres DL, Drummond AJ, Rambaut A. Bayesian phylogenetic and phylodynamic data integration using BEAST 1.10. Virus Evol. 2018 Jun 8;4(1):vey016. doi: 10.1093/ve/vey016

15. Abramson, J., Adler, J., Dunger, J. et al. Accurate structure prediction of biomolecular interactions with AlphaFold 3. Nature 630, 493–500 (2024). 10.1038/s41586-024-07487-w

16. Emsley P, Lohkamp B, Scott WG, Cowtan K. Features and development of Coot. Acta Crystallogr D Biol Crystallogr. 2010 Apr;66(Pt 4):486–501. doi: 10.1107/S0907444910007493. Epub 2010 Mar 24. PMID: 20383002; PMCID: PMC2852313.

17. The PyMOL Molecular Graphics System, Version 3.0 Schrödinger, LLC

18. Papageorgiou N, Coutard B, Lantez V, et al. The 2C putative helicase of echovirus 30 adopts a hexameric ring-shaped structure. Biol Crystallography 2010;66:1116–1120. 10.1107/S090744491002809X.

19. Fratty IS, Kriger O, Weiss L, Vasserman R, Erster O, Mendelson E, Sofer D, Weil M. Increased detection of Echovirus 6-associated meningitis in patients hospitalized during the COVID-19 pandemic, Israel 2021-2022. J. Clin. Vir. 2023;162:105425. Doi.org/10.1016/j.jec.2023.105425.

20. HRSA National Center for Health Workforce Analysis, Health Workforce Projections: Health Technologist and Technician Occupations. https://bhw.hrsa.gov/sites/default/files/bhw/nchwa/projections/healthtechnologisttechniciansapril2015.pdf, accessed February 23, 2018.

21. Zhang M, Xu D, Liu Y, Wang X, Xu L, Gao N, Feng C, Guo W, Ma S. Screening of a new candidate coxsackievirus B1 vaccine strain based on its biological characteristics. Front. Microbiol. 2023;14:1172349.

22. Yeager C, Carter G, Gohara DW, et al. Enteroviral 2C protein is an RNA-stimulated ATPase and uses a two-step mechanism for binding to RNA and ATP. Nucleic Acids Research 2022;50:11775–11798.

